# Modulate Obesity and relateD metabolic complIcations For Yielding improvements in IBD outcomes (MODIFY-IBD): Consensus on Obesity and Cardiometabolic Comorbidities in Inflammatory Bowel Disease using Evidence Synthesis and the RAND/UCLA Appropriateness Method

**DOI:** 10.64898/2025.12.20.25342738

**Authors:** Jalpa Devi, Sami Samaan, Priya Sehgal, Mouhand Mohamed, Matthew Vincent, Shannon Coombs, Michelle Doering, Edward L. Barnes, Amanda M. Johnson, Andres J. Yarur, Parakkal Deepak

## Abstract

**Introduction:** Obesity and related cardiometabolic comorbidities, including hypertension, dyslipidemia, diabetes, metabolic dysfunction-associated steatotic liver disease (MASLD), and atherosclerotic cardiovascular disease (ASCVD), are increasingly prevalent among individuals with inflammatory bowel disease (IBD). These conditions influence disease activity, therapeutic response, surgical outcomes, and overall quality of life, yet evidence remains fragmented. The **M**odulate **O**besity and relate**D** metabolic compl**I**cations **F**or **Y**ielding improvements in **IBD** outcomes (MODIFY-IBD) initiative aims to synthesize evidence and generate consensus recommendations to guide practice and future research in this area.

**Methods and analysis:** We will conduct a structured evidence review organized into three domains: (1) the impact of obesity on IBD outcomes (2) burden of cardiometabolic complications in IBD, and (3) management of obesity and cardiometabolic comorbidities in IBD. Draft clinical statements will be generated and evidence summaries prepared using the GRADE (Grading of Recommendations, Assessment, Development and Evaluation) framework, with certainty of evidence rated where applicable. In the assessment of those statements where GRADE is not feasible, a multidisciplinary international panel of gastroenterologists, surgeons, endocrinologists, hepatologists, cardiologists, and dietitians will assess each statement using the RAND/UCLA Appropriateness Method. Panelists will rate the appropriateness of each statement (only those that fall into their area of expertise) on a 1-9 scale (1-3 = inappropriate, 4-6 = uncertain, 7-9 = appropriate), with medians rounded up (e.g., 6.5 = appropriate). Agreement will be assessed using the RAND Disagreement Index (DI <1.0 = agreement).

**Ethics and dissemination:** This study will not involve direct patient participation, as it is based on evidence synthesis and expert consensus; therefore, formal Research Ethics Committee approval will not be required. Patient representatives will contribute to the consensus process to provide contextual perspectives, but no identifiable data will be collected.

Findings will be disseminated through publication in peer-reviewed journals, presentation at major gastroenterology and IBD conferences, and communication with professional societies. A lay summary and patient-friendly infographic will also be developed to facilitate translation of recommendations into clinical practice.

**PROSPERO registration number:** CRD420251178843: A systematic review of the impact of obesity on inflammatory bowel disease outcomes

CRD420251178799: A Systematic Review of Cardiometabolic Complications in Inflammatory Bowel Disease

CRD420251174653: Management of Overweight, Obesity, and Cardiometabolic Comorbidities in Inflammatory Bowel Disease: A Systematic Review

**Strengths and limitations:**

- Integrates systematic evidence synthesis with both GRADE and RAND/UCLA methods, an approach not previously applied to obesity and cardiometabolic comorbidities in IBD.
- International, multidisciplinary panel (gastroenterology, surgery, endocrinology, cardiology, dietetics, radiology) ensures broad expertise.
- Anonymous scoring and iterative re-rating reduce bias while enabling structured discussion.
- Patient and public involvement will inform priorities and dissemination, although RAND scoring remains expert-only.
- Evidence is largely observational and rapidly evolving (e.g., Glucagon-Like Peptide-1 Receptor Agonists [GLP-1Ras], endobariatric therapies), necessitating future updates.

## Introduction

Obesity and cardiometabolic comorbidities are increasingly recognized as important modifiers of disease course and outcomes in inflammatory bowel diseases (IBD). Recent epidemiological studies estimate that 12-40% of adults with IBD are obese, with an additional 25-40% qualifying as overweight(1, 2). The prevalence of metabolic syndrome approaches 19%, with even higher rates seen in ulcerative colitis (UC) than Crohn’s disease (CD), and risks of type 2 diabetes (T2DM), cardiovascular disease (CVD), and metabolic dysfunction-associated steatotic liver disease (MASLD) are elevated compared with the general population(3–7). Obesity is linked to more active and relapsing disease, greater need for hospitalization, and increased extraintestinal manifestations(8, 9). Therapeutic response is attenuated in patients with IBD and obesity, particularly to biologics and immunomodulators, due to altered drug pharmacokinetics and lower trough levels, resulting in reduced rates of steroid-free remission(10, 11). Surgical outcomes are also negatively affected, with higher perioperative complication rates, longer hospital stays, and greater healthcare costs, risks that are most pronounced with excess visceral adiposity(12, 13). Beyond medical and surgical outcomes, obesity and cardiometabolic complications are associated with impaired quality of life, including higher levels of fatigue, anxiety, depression, and pain(9, 14). Moreover, the rapid expansion of anti-obesity therapies, such as glucagon-like peptide-1 receptor agonists (GLP-1RAs), bariatric surgery, and bariatric endoscopy, has outpaced the development of IBD-specific guidance(15–17).

The lack of harmonized definitions and recommendations has led to fragmented care, with variability in practice across regions and specialties. As such, there is an urgent need for a rigorous, evidence-based, and expert-driven consensus to guide clinical care and new discovery.

## Aims & Objectives

The primary objective of the **M**odulate **O**besity and relate**D** metabolic compl**I**cations **F**or **Y**ielding improvements in **IBD** outcomes (MODIFY-IBD) initiative is to develop evidence-based consensus recommendations for the management of obesity and cardiometabolic comorbidities in patients with IBD, informed by systematic review and RAND/UCLA expert consensus methodology.

### Secondary objectives include

- To define and standardize the assessment of obesity and cardiometabolic comorbidities in IBD (e.g., body mass index (BMI), visceral adipose tissue (VAT), body composition, metabolic syndrome, MASLD).
- To evaluate the impact of obesity and metabolic syndrome on medical and surgical outcomes in IBD, including response to biologics, treatment durability, post-operative complications, and disease recurrence.
- To provide guidance on the role and impact of lifestyle interventions, pharmacotherapy, bariatric surgery, and bariatric endoscopy in the management of patients with IBD and obesity.
- To highlight gaps in evidence and set research priorities for future clinical trials and translational studies.

### Clinical domains

Three domains with subtopics will structure the review and consensus process across three work groups:

Domain 1: Management of Overweight, Obesity, and Cardiometabolic Comorbidities in IBD

- Measures and definitions of obesity and visceral adiposity in IBD
- Lifestyle interventions (dietary modifications, physical activity, behavioral interventions)
- Pharmacologic management

- Anti-obesity therapies (including GLP-1RAs)
- Antidiabetic, antihypertensive, and lipid-lowering therapies
- Multidisciplinary care models and preventive strategies
- Bariatric and endobariatric procedures in IBD

Domain 2: Cardiometabolic Complications in Inflammatory Bowel Disease

- Epidemiology of cardiometabolic comorbidities in IBD
- Metabolic dysfunction–associated steatotic liver disease (MASLD/MASH) and its complications in IBD
- Cardiovascular disease and major adverse cardiovascular events in IBD
- Metabolic syndrome, diabetes, hypertension, and dyslipidemia in IBD
- Impact of cardiometabolic complications on IBD-related outcomes

Domain 3: Impact of Obesity on Inflammatory Bowel Disease Outcomes

- besity and disease activity, severity, and natural history
- besity and response to medical therapy
- besity and surgical outcomes in IBD
- besity, body composition, and inflammatory biomarkers
- effects by IBD subtype and adiposity phenotype

## Methods and analysis

### Study design and registration

This consensus project will integrate systematic evidence synthesis and RAND/UCLA expert consensus. The three systematic reviews are registered with PROSPERO (CRD420251174653, CRD420251178799, CRD420251178843).

### Timeline

- Start date: September 1, 2025
- Completion date: November 30, 2026

### Part 1: Systematic Review

#### Study selection and search strategy

The evidence review will be conducted in accordance with PRISMA guidelines and registered with PROSPERO. A comprehensive search of MEDLINE, Embase, Web of Science, and the Cochrane Library (1990–2025) will be undertaken, supplemented with clinical trial registries (ClinicalTrials.gov, EU CTR, ISRCTN, WHO ICTRP) and grey literature. Search strategies will use controlled vocabulary and free-text terms for inflammatory bowel disease, obesity, overweight, weight management, cardiometabolic conditions, and relevant interventions (lifestyle, pharmacologic, surgical, endobariatric). Inclusion and exclusion criteria will be applied to identify observational and interventional studies in adults with IBD. (**Supplementary Appendix 1**: Search Strategies, Data Extraction Framework, and Planned Data Synthesis for the Three Systematic Reviews (MODIFY-IBD Initiative))

#### Data extraction

Two independent reviewers will screen titles and abstracts in Covidence (Veritas Health Innovation) for each of the three workgroups. A predefined extraction form will capture study design, population characteristics, interventions, comparators, outcomes, and adverse events. For pharmacotherapy, dose and duration will be recorded; for lifestyle interventions, type, frequency, and intensity; for surgical approaches, procedure type and peri-operative details. Discrepancies will be resolved by discussion with a third independent reviewer.

#### Risk-of-bias (quality) assessment

Risk of bias will be assessed independently by two reviewers. The Cochrane Risk of Bias 2 tool will be used for randomized controlled trials, and ROBINS-I for observational studies. Methodological quality will be tabulated, and any disagreements adjudicated by consensus.

#### Strategy for data synthesis

Heterogeneity in study design and outcome measures is anticipated. Where feasible, random-effects meta-analysis will be performed, reporting mean differences (continuous outcomes) or risk ratios (RR)/odds ratios (OR) (dichotomous outcomes) with 95% confidence intervals (CI). Statistical heterogeneity will be assessed using the I² statistic. When meta-analysis is not feasible, a structured narrative synthesis will be provided.

#### Analysis of subgroups or subsets

Planned subgroup analyses include IBD subtype (Crohn’s disease vs ulcerative colitis), BMI category (overweight vs obese), intervention type (lifestyle, pharmacotherapy, psychotherapy, surgical, endobariatric), cardiometabolic comorbidity type (e.g., diabetes, MASLD, ASCVD), and concomitant IBD therapies (biologic vs non-biologic).

### Part 2 Consensus Development Process

#### Selection and recruitment of stakeholders (**Figure 1**)

A multidisciplinary expert panel (target n = 50-60) will be convened, including gastroenterologists, colorectal surgeons, bariatric surgeons, endobariatric endoscopists, endocrinologists, hepatologists, cardiologists, radiologists, dietitians, and pharmacists. Experts will be identified through structured searches of faculty profiles, bibliometric analyses of obesity-IBD publications, and professional society networks (IOIBD, ECCO, AGA, ACG, APAGE). Invitations will be distributed via personalized email from the study lead, detailing study aims, RAND/UCLA process, and expected time commitment (∼20-30 min per round). Eligibility will require demonstrable clinical or research expertise in obesity, metabolic disease, or IBD, with attention to gender, geographic, and career-stage diversity. Participation will be voluntary with electronic consent, and panelists will receive acknowledgment under MODIFY-IBD authorship.

**Figure 1:**
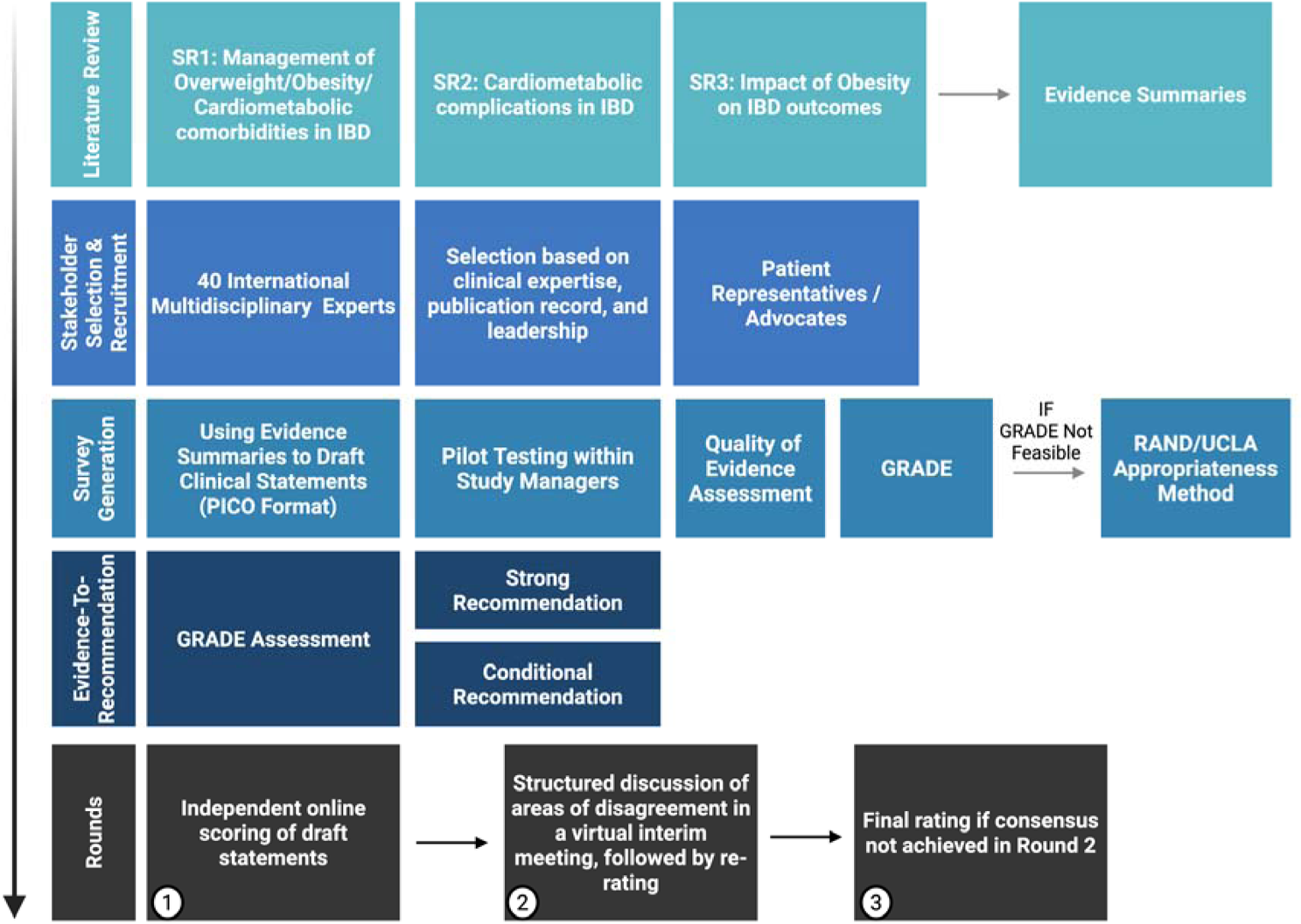
Workflow of the MODIFY-IBD Evidence Synthesis and Consensus Development Process **Created in BioRender. Deepak, P. (2026)** https://BioRender.com/jrd3pyw

Engagement will be promoted through reminder emails, and feedback summaries after each round. Those unable to complete a round will be invited to participate in subsequent rounds to mitigate attrition bias.

#### Rounds description

- **Round 1**: Independent online scoring of draft statements.
- **Round 2**: Structured discussion of areas of disagreement in a virtual interim meeting, followed by re-rating.
- **Round 3**: Final rating if consensus not achieved in Round 2.

#### Data management

Data collected through each survey round will be analysed in aggregate by the study investigators. Washington University in St. Louis will serve as the data custodian, with the lead investigator (PD) responsible for ensuring compliance with institutional privacy and data security standards. All data will be stored on encrypted, access-restricted institutional servers (e.g., REDCap), accessible only to the core research team. No identifiable information will be shared outside the study team. De-identified datasets and summary statistics will be archived for five years following study completion in accordance with institutional policy.

#### Definitions

Obesity will be defined primarily as BMI ≥30 kg/m², with additional body composition metrics (waist circumference, visceral adipose tissue, sarcopenia indices) included where available. Cardiometabolic comorbidities include hypertension, dyslipidemia, type 2 diabetes, metabolic syndrome [(National Cholesterol Education Program-Adult Treatment Panel III) (NCEP-ATP III)] / International Diabetes Federation (IDF)]), atherosclerotic cardiovascular disease, and (Metabolic dysfunction-Associated Steatotic Liver Disease / Metabolic dysfunction-Associated Steatohepatitis) MASLD/MASH.

#### Survey generation

Draft statements will be derived from the systematic review evidence summaries and structured into PICO format where possible. Each statement will undergo pilot testing by the study management group before distribution. Given the paucity of robust data, we will also use the RAND/UCLA appropriateness method, which combines systematic evidence appraisal with expert judgment to generate clinically relevant recommendations.

#### Certainty of Evidence

We will assess the quality of evidence using the GRADE framework (18). RCTs will begin as high-certainty and observational studies as low, with ratings adjusted for bias, inconsistency, imprecision, indirectness, or publication bias. In some cases, observational evidence may be rated up (e.g., large effect, dose-response)(19, 20).

#### From Evidence to Recommendations

We will use the GRADE Evidence-to-Decision framework to weigh benefits, harms, patient values, feasibility, resource use, and equity. Recommendations will be labeled *strong* (“we recommend”) or *conditional* (“we suggest”), with corresponding certainty ratings (**Table 1 & 2**). Implementation guidance will be provided to support clinical adoption(21).

**Table 1:** Interpretation of Strong and Conditional Recommendations Using the GRADE Framework.

| **Implications** | **Strong Recommendation** | **Conditional Recommendation** |
| --- | --- | --- |
| **For patients** | Most individuals in this situation will prefer the recommended course of action; only a small proportion would not. | Many individuals would want the suggested course of action, but a substantial number may choose differently based on personal preferences. |
| **For clinicians** | Most patients should receive the recommended intervention.<br>Formal decision aids are generally not needed. | Shared decision-making is essential.<br>Clinicians should discuss the pros, cons, and alternatives with patients to support decisions aligned with values and preferences. |
| **For policy makers** | The recommendation can be adopted as a policy or performance measure in most settings. | Policymaking requires consideration of diverse stakeholder perspectives.<br>Performance measures should assess decision-making quality, not just adherence. |

**Table 2:** Interpretation of Quality of Evidence (GRADE Framework)

| **Quality of Evidence** | **Definition** |
| --- | --- |
| **High** | We are very confident that the true effect is close to the estimated effect. |
| **Moderate** | We are moderately confident in the effect estimate; the true effect is likely close but may be substantially different. |
| **Low** | Our confidence in the effect estimate is limited; the true effect may be substantially different from the estimate. |
| **Very Low** | We have very little confidence in the effect estimate; the true effect is likely substantially different. |

#### RAND/UCLA Appropriateness Method (RUAM)

To systematically determine the appropriateness of key clinical statements, we will employ the **RAND/UCLA Appropriateness Method (RUAM)**, a validated and widely used modified Delphi technique(22). This method integrates the best available evidence with expert clinical judgment to assess the balance of benefits and harms for each proposed statement; particularly when randomized trial data are limited or absent(23).

The RUAM has demonstrated strong reliability, internal consistency, and clinical validity across a range of medical and surgical disciplines, including IBD(24–28). The RAND/UCLA process will be applied only to questions or statements where formal GRADE assessment is not feasible or where the evidence base is limited.

Each statement will be rated for appropriateness on a 1-9 scale (1-3 = inappropriate, 4-6 = uncertain, 7-9 = appropriate). Median scores will be calculated and rounded up; for example, a median of 3.5 will be classified as uncertain, while a median of 6.5 will be rated as appropriate. Final appropriateness ratings will be based on median scores of 7, 8, or 9.

EQUATION

DI = (66 percentile – 33 percentile) / [2.35 + 1.5 × ((66 percentile + 33 percentile) / 2)]

The measure of agreement will be determined using the RAND Disagreement Index (DI), which incorporates the spread between the 30th and 70th percentiles. A DI ≥ 1.0 indicates disagreement (extreme variation), whereas DI < 1.0 reflects general agreement. Ratings will be conducted in REDCap. Median, interquartile range (IQR), and percentage agreement will be calculated per item. Indicators rated 7-9 by ≥ 50 % of participants in Round 1 will advance to Round 2. Consensus will be defined as median 7-9 and RAND Disagreement Index < 1.0; change in ratings between rounds will be assessed using Wilcoxon signed-rank test.

#### Data analysis

Quantitative and qualitative data will be analysed concurrently. For each statement, we will report the median, interquartile range, and percentage agreement to describe the level of consensus. Changes in ratings between rounds will be assessed using the Wilcoxon signed-rank test. If response bias is suspected, subgroup analyses by specialty or region will be performed. Free-text responses will undergo thematic analysis to identify common viewpoints or reasons for disagreement. In subsequent rounds, participants will receive anonymised feedback comparing their scores with group summaries to support reflection and re-rating.

#### Definition of study completion

The study will be considered complete once the final consensus meeting and last round of re-rating are concluded. At this stage, no new data will be collected, and results will be synthesized into recommendations.

#### Patient and public involvement

Patient representatives will be embedded within the MODIFY-IBD study management group to ensure outcomes remain relevant, accessible, and patient-centered. While patients will not directly participate in the RAND/UCLA rating process due to the technical nature of the questions, they will help guide project priorities, provide feedback on statement wording, and co-develop dissemination materials tailored for the wider IBD community.

#### Rigour and limitations

As with all consensus studies, potential biases may affect validity and reliability(29). We will reduce selection bias by applying predefined eligibility criteria and ensuring multidisciplinary, geographically diverse representation rather than relying solely on personal networks(30). To limit attrition, surveys will be restricted to two to three rounds with reminders(31). Anonymised scoring after group discussion will help prevent groupthink(32). Transparent reporting, protocol registration, and iterative feedback from panelists will ensure methodological rigour and reproducibility(29).

### Ethics and dissemination

#### Risks and consent

No foreseeable risks are expected for participants. Informed electronic consent will be obtained before participation, and panelists may withdraw at any time without consequence.

#### Confidentiality

Responses will be anonymised and linked to unique IDs. Only aggregated data will be shared for group feedback, and all files will be stored on secure, institution-approved server of Redcap of Washington University in Saint Louis, accessible to the study team.

#### Ethics approval

As this study involves evidence synthesis and expert consensus without personal data, formal ethics board approval is not required.

#### Dissemination

Results will be shared through peer-reviewed publications, conference presentations, and summaries for professional societies and patient groups. A lay infographic and open-access supplementary materials will accompany the final publication.

## Discussion

This study will generate a structured, evidence-based framework to guide the management of obesity and related cardiometabolic comorbidities in patients with IBD. By combining systematic evidence synthesis with the RUAM, the MODIFY-IBD initiative will provide a measurable, consensus-driven set of statements to inform clinical decision-making, determine research priorities, and support guideline development.

While the consensus process is grounded in published data, certain knowledge gaps remain due to limited high-quality studies specific to IBD populations. For example, evidence guiding optimal use of GLP-1 RAs, endobariatric procedures, and multidisciplinary care pathways in IBD is still evolving. Similarly, real-world data on long-term cardiometabolic outcomes, such as major adverse cardiovascular events or MASLD progression, are sparse. These limitations highlight the importance of expert input and transparent reporting of uncertainty through GRADE and RAND/UCLA methodologies.

Because expert panels are composed of clinicians, researchers, and patient representatives from multiple specialties and regions, some variation in interpretation is expected. However, using structured rating rounds, anonymised scoring, and iterative feedback will mitigate bias and enhance reproducibility.

Ultimately, this work seeks to establish common definitions, treatment paradigms, and evidence-to-decision frameworks that will support high-quality, multidisciplinary care for patients with obesity and IBD. The resulting statements will also inform the design of future clinical trials, quality improvement initiatives, and implementation studies focused on cardiometabolic risk reduction in IBD.

### Progress to date

Preparatory work including literature searches, PROSPERO registrations, and initial panel invitations was completed between September and November 2025. The first round of surveys is planned for early 2026, followed by iterative analysis and a virtual consensus meeting later that year.

## Data Availability

All data used in this study are derived from publicly available sources and published literature. No new datasets were generated or analyzed. All data relevant to the study are included in the manuscript and its supplementary materials.

## Abbreviations

ACG: American College of Gastroenterology
AGA: American Gastroenterological Association
APAGE: Asian Pacific Association of Gastroenterology
ASCVD: Atherosclerotic Cardiovascular Disease
BMI: Body Mass Index
CD: Crohn’s Disease
CI: Confidence Interval
DDRCC: Digestive Diseases Research Core Center
DI: Disagreement Index
ECCO: European Crohn’s and Colitis Organisation
EtD: Evidence-to-Decision
EU CTR: European Union Clinical Trials Register
GLP-1RA: Glucagon-Like Peptide-1 Receptor Agonist
GRADE: Grading of Recommendations Assessment, Development and Evaluation
IBD: Inflammatory Bowel Disease
IDF: International Diabetes Federation
IOIBD: International Organization for the Study of Inflammatory Bowel Disease
IQR: Interquartile Range
ISRCTN: International Standard Randomised Controlled Trial Number
MASLD: Metabolic Dysfunction-Associated Steatotic Liver Disease
MASH: Metabolic Dysfunction-Associated Steatohepatitis
MD: Mean Difference
MODIFY-IBD: Modulate Obesity and relateD metabolic complIcations For Yielding improvements in IBD outcomes
NCEP-ATP III: National Cholesterol Education Program Adult Treatment Panel III
NIDDK: National Institute of Diabetes and Digestive and Kidney Diseases
OR: Odds Ratio
PICO: Population, Intervention, Comparator, Outcome
PRISMA: Preferred Reporting Items for Systematic Reviews and Meta-Analyses
PROSPERO: International Prospective Register of Systematic Reviews
RCT: Randomized Controlled Trial
REDCap: Research Electronic Data Capture
ROBINS-I: Risk Of Bias In Non-randomized Studies of Interventions
RR: Risk Ratio
RUAM: RAND/UCLA Appropriateness Method
SR: Systematic Review
UC: Ulcerative Colitis
VAT: Visceral Adipose Tissue
WHO ICTRP: World Health Organization International Clinical Trials Registry Platform

