## Supplementary material for "Modulate Obesity and relateD metabolic complIcations For Yielding improvements in IBD outcomes (MODIFY-IBD): Consensus on Obesity and Cardiometabolic Comorbidities in Inflammatory Bowel Disease using Evidence Synthesis and the RAND/UCLA Appropriateness Method": Supplemetary file

**Supplementary Appendix 1: Search Strategy for Literature Review**

Three systematic literature reviews were conducted in collaboration with an expert librarian at Washington University in St Louis Becker Library. Two reviewers from the study managers were assigned for each review, with a third reviewer for conflict management during the screening stage.

Risk of bias will be assessed independently by two reviewers. For randomized controlled trials, the Cochrane Risk of Bias 2 tool will be used; for observational studies, the ROBINS-I tool will be applied. Methodological quality, reporting transparency, and risk of bias across key domains (selection, performance, detection, attrition, reporting) will be evaluated.

**1^st^ Systematic Review: Management of Overweight/Obesity/Cardiometabolic comorbidities in IBD**

Medical literature databases were searched using strategies created by a librarian using a combination of standardized terms and keywords for inflammatory bowel diseases, ulcerative colitis, and Crohn disease combined with terms for obesity, overweight, visceral adipose tissue, type 2 diabetes, hypertension, hyperlipidemia, atherosclerosis, and non-alcoholic fatty liver disease combined with terms for diet, exercise, weight-lowering medications, bariatric surgery, antidiabetic agents, lipid-lowering agents, antiplatelet agents, and antihypertensives. The search was run in the databases Embase.com, Ovid Medline, PubMed, Scopus, Cochrane Central, and Clinicaltrials.gov on September 2, 2025. The search included a date limit for 1991 to current, an English language limit, a librarian-created filter to exclude pediatric studies, and a librarian-created filter to exclude animal studies. A total of 7,752 citations were imported into Covidence. 2,785 duplicates were assumed to be accurately identified and removed for a total of 4,967 unique citations for screening.

**Supplementary Table 1**: PICO for Management of Overweight/Obesity/Cardiometabolic comorbidities in IBD

| Condition Studied | Weight management strategies in IBD including lifestyle, interventions, pharmacotherapy, endobariatric procedures, and surgical approaches |
| --- | --- |
| Population | The population of patients included in analysis will be:  • Adults (≥18 years) with a diagnosis of Crohn’s disease or ulcerative colitis who are overweight or obese (BMI ≥25 kg/m²).  The following patients will be excluded:  • Pediatric populations (<18 years)  • Animal studies |
| Intervention(s) or exposure(s) | Any intervention aimed at managing overweight or obesity, or associated cardiometabolic disorders, including:  • Structured lifestyle interventions (diet, physical activity, behavioral therapy)  • Pharmacotherapy (GLP-1 receptor agonists, other anti-obesity medications, antihypertensives, lipid-lowering therapy, glycemic control agents)  • Bariatric surgery  • Endobariatric procedures |
| Comparator(s) or control(s) | • IBD patients receiving no specific weight-loss intervention, usual care, or alternative strategies  • Non-IBD patients undergoing similar interventions |
| Outcomes | **Main outcomes**  The primary outcome of this systematic review will be to assess the effectiveness of interventions for weight management in adults with IBD who are overweight or obese, measured by:  • Absolute weight loss (kg)  • Percentage weight loss (%)  • Change in body mass index (BMI)  Comparisons will be made with IBD patients receiving usual care/alternative interventions, and with non-IBD patients undergoing similar interventions.  **Measures of effect**  We will report outcomes as mean difference (MD) or standardized mean difference (SMD) for continuous variables, and risk ratios (RR) or odds ratios (OR) for dichotomous variables, with 95% confidence intervals. Where meta-analysis is not feasible, a narrative synthesis will be provided.  Additional outcomes  Secondary outcomes will include:  • Safety: Adverse events and serious adverse events, comparing IBD with non-IBD populations.  • IBD-specific outcomes: Clinical, endoscopic, or radiological remission; relapses; corticosteroid use; need for escalation of therapy; IBD-related surgery or hospitalization.  • Biomarkers: Change in C-reactive protein (CRP) and fecal calprotectin.  • Quality of life: Assessed using validated patient-reported outcome measures.  • Cardiometabolic outcomes: Change in blood pressure, lipo-protein profile, glycemic control (HbA1c, insulin resistance indices), MACE (MI, stroke, cardiovascular death, heart failure), and presence or progression of MASLD/MASH. |

**Data Extraction**

A predefined data extraction form will be designed and implemented in Covidence. Extracted data will include study characteristics, design and methodology, patient demographics, baseline characteristics, intervention type, comparator, and primary and secondary outcomes. For pharmacotherapy trials, specific drug, dose, and duration will be recorded; for lifestyle interventions, type, frequency, and intensity will be noted; and for surgical interventions, type and peri-operative details will be captured. Cardiometabolic outcomes (e.g., blood pressure, lipids, HbA1c, MASLD/MASH markers) will also be extracted where available. Two reviewers will independently extract data, and discrepancies will be resolved through discussion with a senior author. Attempts will be made to contact study authors for clarification or missing data.

**PLANNED DATA SYNTHESIS**

Strategy for data synthesis

Given the expected heterogeneity in study populations, interventions, and outcomes, a narrative synthesis will be undertaken where quantitative pooling is not feasible. Where studies are sufficiently homogeneous in design and reporting, meta-analysis will be performed using a random-effects model. Continuous outcomes (e.g., weight change, BMI, metabolic parameters) will be expressed as mean difference (MD) or standardized mean difference (SMD), and dichotomous outcomes (e.g., remission, adverse events) as risk ratios (RR) or odds ratios (OR), each with 95% confidence intervals. Heterogeneity will be assessed using the I² statistic. If meta-analysis is not possible, findings will be reported descriptively. The results of data extraction and methodological quality assessment will be synthesized narratively in the final publication.

Analysis of subgroups or subsets

Planned subgroup analyses will include:

•	IBD subtype: Crohn’s disease vs ulcerative colitis

•	Type of intervention: lifestyle (diet, exercise), pharmacotherapy (GLP-1 receptor agonists and other anti-obesity drugs, cardiometabolic agents), bariatric surgery, endobariatric procedures

•	Comparator group: IBD vs non-IBD populations undergoing the same intervention

•	Baseline BMI categories: overweight (25-29.9 kg/m²) vs obese (≥30 kg/m²)

•	Concomitant IBD therapies: advanced therapy exposure vs advanced therapy–naïve

Where sufficient data are available, we will also explore the effect of interventions on cardiometabolic outcomes (blood pressure, HbA1c, lipid profile, MASLD/MASH markers) as subgroup analyses.

**2^nd^ Systematic Review: Cardiometabolic complications in IBD**

Medical literature databases were searched using strategies created by a librarian using a combination of standardized terms and keywords for inflammatory bowel diseases, ulcerative colitis, and Crohn disease combined with terms for cardiometabolic disease, obesity, type 2 diabetes, cardiovascular disease, and MASLD combined with terms for risk, association, link, prevalence, and incidence combined with terms for compared, non-IBD, without-IBD, cohort, controls, retrospective, observational, or matched. The search was run in the databases Embase.com, Ovid Medline, PubMed, Scopus, and Cochrane Central on September 11, 2025. The search included a date limit for 1991 to current, an English language limit, a librarian-created filter to exclude pediatric studies, and a librarian-created filter to exclude animal studies. A total of 13,269 citations were imported into Covidence. 5,008 duplicates were assumed to be accurately identified and removed for a total of 8,261 unique citations for screening.

**Supplementary table 2**: PICO for Cardiometabolic complications in IBD

| Condition Studied | Inflammatory Bowel Disease and Cardiometabolic Conditions |
| --- | --- |
| Population | The population of patients included in analysis will be:  • Adults (≥18 years) with a diagnosis of Crohn’s disease or ulcerative colitis  The following patients will be excluded:  • Pediatric populations (<18 years)  • Animal studies |
| Intervention(s) or exposure(s) | Presence of IBD (as an exposure variable); stratified where applicable by disease subtype, duration, activity, and treatment class |
| Comparator(s) or control(s) | Individuals without IBD (general population controls AND without other chronic inflammatory diseases (e.g., rheumatoid arthritis, psoriasis, psoriatic arthropathy) |
| Outcomes | **Main outcomes**  • Incidence and/or prevalence of cardiometabolic conditions, including:  • Cardiovascular disease (e.g., myocardial infarction, stroke, heart failure)  • Type 2 diabetes mellitus  • Hypertension  • Dyslipidemia  • Metabolic syndrome  • MASLD/MASH  • Overweight/obesity  **Additional outcomes**  • Subclinical markers (e.g., carotid intima-media thickness, coronary artery calcium score, arterial stiffness)  • Insulin resistance and hyperglycemia  • All-cause and cardiovascular-specific mortality |

**Data Extraction**

A predefined data extraction form will be designed and implemented using Covidence.

The target condition of interest is inflammatory bowel disease. Extracted data will include study characteristics, design and methodology, patient demographics, and baseline characteristics. Primary and secondary outcomes will be recorded. 2 reviewers will independently extract data and conflicts will be resolved through discussion with senior authors. Attempts will be made to obtain missing data through contacting senior authors of studies included for review.

**PLANNED DATA SYNTHESIS**

Strategy for data synthesis

A narrative synthesis will be conducted to summarize the incidence and prevalence of cardiometabolic complications in individuals with IBD compared to non-IBD controls. Where data are sufficiently homogeneous in terms of study design, population characteristics, outcome definitions, and measurement methods, a meta-analysis will be performed using a random-effects model to account for between-study variability. Pooled estimates (risk ratios, odds ratios, or incidence rate) will be calculated along with corresponding 95% confidence intervals. Heterogeneity will be assessed using the I² statistic, with values >50% indicating substantial heterogeneity. Sensitivity analyses will be performed to evaluate the impact of study quality (based on risk of bias assessment) and study design. Publication bias will be assessed using funnel plots.

Analysis of subgroups or subsets

Subgroup analyses will be conducted based on IBD subtype (Crohn’s disease vs. ulcerative colitis), disease activity, treatment exposure, and study setting if sufficient data are available.

**3^rd^ Systematic Review: Impact of Obesity on IBD outcomes**

The published literature was searched using strategies created by a medical librarian using a combination of standardized terms and keywords for inflammatory bowel diseases, ulcerative colitis, Crohn disease, obesity, overweight, visceral adipose tissue, complications, outcomes, treatment response, remission, infection, case control studies, retrospective studies, comparison groups, and non-obese controls. The search was run in the databases Embase.com, Ovid Medline, PubMed, Scopus, and Cochrane Central on August 18, 2025. The search included a date limit for 1991 to current, an English language limit, a librarian-created filter to exclude pediatric studies, and a librarian-created filter to exclude animal studies. A total of 3,134 citations were imported into Covidence. 585 duplicates were assumed to be accurately identified and removed for a total of 2,549 unique citations for screening.

Supplementary table 3: PICO for Impact of Obesity on IBD outcomes

| Condition Studied | Obesity effects on both the disease course and response to interventions in the adult IBD population |
| --- | --- |
| Population | The population of patients included in analysis will be:  • Adults (≥18 years) with a diagnosis of Crohn’s disease or ulcerative colitis  • Obese group: BMI ≥30 kg/m² or other measures of adiposity (waist circumference, visceral adiposity, body composition imaging-based measures).  The following patients will be excluded:  • Pediatric populations (<18 years)  • Animal studies |
| Intervention(s) or exposure(s) | The primary exposure evaluated will be obesity, defined as body mass index (BMI) ≥30 kg/m². Because BMI does not fully capture variations in body fat distribution or metabolic risk, we will also evaluate obesity utilizing alternate measures of adiposity. These alternate measures will include visceral adiposity (VAT) or visceral-to-subcutaneous adipose tissue ratio (VAT:SAT) (VFI), quantified where available by imaging (including computed tomography [CT] scan, dual energy x-ray absorptiometry [DEXA] scan, InBody Scales) as well as the waist-to-hip ratio |
| Comparator(s) or control(s) | Non-obese individuals with IBD (BMI <30 kg/m²). Given we are including other surrogate measures of obesity to include VAT, VFI, and waist-to-hip ratio, we will compare patients with obesity based on these metrics to their “non-obese” counterparts within those respective definitions. |
| Outcomes | **Primary Outcome:**   - Impact of obesity on clinical and endoscopic remission in patients with inflammatory bowel disease (IBD). Clinical remission will be defined according to various validated disease activity indices (including Crohn’s disease activity index [CDAI], Harvey Bradshaw Index [HBI], Mayo scores) whereas endoscopic remission will be defined by endoscopic indices (simple endoscopic score for Crohn’s disease [SES-CD], Mayo endoscopic sub-score).   **Additional outcomes:**   - Need for oral or IV steroids, need to escalate or change in advanced therapy, therapeutic drug monitoring, IBD-related hospitalization, IBD-related surgery, post-operative disease recurrence (CD), quality of life, mortality (all-cause and IBD-related). - Post-operative complications (wound or intra-abdominal infection, anastomotic leak, VTE, unplanned stoma), technical feasibility of IPAA, re-admission post-surgery, re-operation rates, length of hospital stay, open vs laparoscopic surgery rates, stoma-related complications, surgical recurrence (especially in Crohn’s disease), IPAA-associated complications including pouchitis, pouch failure rates (pouch excision, diverting loop ileostomy). |

**Data extraction**

A predefined data extraction form will be designed and implemented using Covidence. Extracted data will include, but are not limited to: study characteristics (study design, setting, study period, sample size, source of funding, inclusion/exclusion criteria, geographic region); population characteristics (IBD type, age, sex distribution, BMI/obesity category, IBD duration, disease activity at baseline, medication exposure, baseline cardiometabolic characteristics; baseline BMI/adiposity measures (visceral adiposity, VAT:SAT, waist to hip ratio, or other body composition imaging), medical therapy details, surgical type, and outcomes. At least two study authors (PS and MM) will abstract the data from the included studies into preprepared data collection sheets.

**PLANNED DATA SYNTHESIS**

Strategy for data synthesis

When quantitative analysis is not feasible due to heterogeneity in study design, outcome definitions, we will perform a narrative synthesis of the evidence. Narrative synthesis will summarize findings thematically across study types, highlighting consistencies, divergences and gaps in literature. Particular attention will be paid to differences in outcome definitions and obesity/adiposity measurements thay may account for heterogeneity.

Where sufficient data are available, a meta-analysis using a random-effects model will be conducted to account for between study variability. Heterogeneity will be assessed via the I² statistic.

Analysis of subgroups or subsets

To better understand the relationship between obesity and outcomes in IBD, we will conduct several different subgroup analyses. We will evaluate differences in outcomes of patients with elevated BMI versus lower BMI, and between those with elevated visceral adipose tissue vs lower visceral adipose tissue (VAT). When possible, we will also compare BMI-based measures with volumetric analysis as well.

We will also assess whether obesity impacts response to advanced therapies, need for steroids, and need for surgical intervention.

Supplementary Table 4. Recommendations for the Conducting and Reporting of Delphi Studies (CREDES) Checklist

| Reporting Items | Reported on page |
| --- | --- |
| *Purpose and rationale.* The purpose of the study should be clearly defined and  demonstrate the appropriateness of the use of the Delphi technique as a  method to achieve the research aim. A rationale for the choice of the Delphi  technique as the most suitable method needs to be provided. | 1-3 |
| *Expert panel.* Criteria for the selection of experts and transparent information on  recruitment of the expert panel, sociodemographic details including information  on expertise regarding the topic in question, (non)response and response rates  over the ongoing iterations should be reported. | 5 |
| *Description of the methods.* The methods employed need to be comprehensible.  this includes information on preparatory steps (How was available evidence on  the topic in question synthesized?), piloting of material and survey instruments,  design of the survey instrument(s), the number and design of survey rounds,  methods of data analysis, processing and synthesis of experts’ responses to  inform the subsequent survey round and methodological decisions taken by the  research team throughout the process. | 4-7 |
| *Procedure.* Flow chart to illustrate the stages of the Delphi process,  including a preparatory phase, the actual ‘Delphi rounds’, interim steps of  data processing and analysis and concluding steps. | Figure1 |
| *Definition and attainment of consensus.* It needs to be comprehensible to the  reader how consensus was achieved throughout the process, including  strategies to deal with non-consensus. | 8 |
| *Results.* Reporting of results for each round separately is highly advisable in  order to make the evolving of consensus over the rounds transparent. This  includes figures showing the average group response, changes between  rounds, as well as any modifications of the survey instrument such as deletion,  addition or modification of survey items based on previous rounds. | To be added at the end of the study |
| *Discussion of limitations.* Reporting should include a critical reflection of potential  limitations and their impact of the resulting guidance. | 2 |
| *Adequacy of conclusions.* The conclusions should adequately reflect the  outcomes of the Delphi study with a view to the scope and applicability of the  resulting practice guidance. | To be added at the end of the study |
| *Publication and dissemination.* The resulting guidance on good practice in  palliative care should be clearly identifiable from the publication, including  recommendations for transfer into practice and implementation. If the  publication does not allow for a detailed presentation of either the resulting  practice guidance or the methodological features of the applied Delphi  technique, or both, reference to a more detailed presentation elsewhere  should be made (e.g. availability of the full guideline from the authors or  online, publication of a separate paper reporting on methodological details and  particularities of the process (e.g. persistent disagreement and controversy on  certain issues). A dissemination plan should include endorsement of the  guidance by professional associations and health care authorities to facilitate  implementation. | 8 |

The RAND/UCLA method follows modified Delphi principles; hence, CREDES items were used as a guide for transparent reporting.

Jünger, S., Payne, S. A., Brine, J., Radbruch, L., & Brearley, S. G. (2017). Guidance on Conducting and

REporting DElphi Studies (CREDES) in palliative care: Recommendations based on a methodological systematic

review. Palliative medicine, 31(8), 684-706.
